# The national burden from O’nyong’nyong and chikungunya viruses in Cameroon

**DOI:** 10.64898/2026.06.24.26356335

**Authors:** Ngu Njei Abanda, Anchita Puri, Michael White, Laura Garcia, Camille Lambert, Sorel Jakpou, Jules Brice Tchatchueng Mbougua, Oscar Cortes-Azuero, Scott C. Weaver, Henrik Salje, Richard Njouom

## Abstract

O’nyong-nyong (ONNV) and chikungunya (CHIKV) are arboviruses that have been identified nearly throughout Africa. However, the disease burden from the viruses has been rarely quantified as surveillance systems are poorly equipped to identify cases. Here, we tested 6,324 serum samples collected as part of the Yellow Fever testing program between 2010 and 2023 in individuals of all ages from all the health districts of Cameroon. We used a novel multiplex assay that incorporated antigens from three alphaviruses (ONNV, CHIKV and MAYV) and mathematical models that explicitly consider cross-reactivity to reconstruct individual and population infection histories for each virus. We found that despite no cases reported in Cameroon, ONNV has circulated endemically across the country for decades with an average of 332,000 annual infections and 8 million individuals with a history of infection. By contrast, we found evidence of only sporadic CHIKV outbreaks, with an estimated prevalence of 1.7%. Models that did not explicitly consider cross-reactivity between the alphaviruses lead to incorrect conclusions about the circulation of the viruses. This work highlights the need for systematic efforts to identify ONNV incident cases and the development of a vaccine development investment case against this neglected pathogen.

## Main

Mosquito transmitted viruses (arboviruses), such as dengue (DENV), Zika (ZIKV) and chikungunya (CHIKV) viruses continue to be a major threat to public health worldwide^1^. However, the true burden caused by these viruses is rarely determined. Lack of appropriate diagnostic kits, limited healthcare seeking in formal healthcare settings and frequent subclinical infection mean that most infections are not captured by surveillance systems and even entire large epidemics can be missed^2^. The knowledge gap is particularly severe for alphaviruses in low-income settings. We know that viruses such as CHIKV, transmitted by *Aedes* mosquitoes, and o’nyong-nyong virus (ONNV), transmitted by *Anopheles*, are present in Africa but the level of transmission is not known, including the extent of endemic versus epidemic transmission and key predictors of infection risk. These alphaviruses are a particular concern to public health due to the potential for long-term arthralgia or death following infection^3^. Understanding which pathogens circulate within a community is central to targeted public health action, including the use of vaccines or non-pharmaceutical interventions as well as vector control.

CHIKV, transmitted to humans during epidemics by infected *Aedes aegypti* and *Ae. albopictus* mosquitoes, is associated with large outbreaks and sporadic cases reported mostly in the Americas, Asia and Africa, and occasional smaller outbreaks in Europe ^4^. Spillover infections from nonhuman primate-amplified sylvatic, enzootic cycles also occur, and it has been suggested that CHIKV is endemic in East Africa ^5^. ONNV, primarily transmitted by *Anopheles funestus* and *An. gambiae* has only been detected in Africa. Whilst large ONNV outbreaks have occurred in the past, with up to 2 million cases reported in East and West Africa between 1959 and 1962, there is very little understanding of its current circulation in Africa, especially its presumed enzootic cycle ^6^.

In this study we focused on Cameroon. The country provides an excellent opportunity to understand potential drivers of CHIKV and ONNV ecology. The North of the country is very hot and touches the Saharan desert, whereas the South is tropical. *Anopheles* and *Aedes* mosquitoes are found throughout the country^7, 8^. To understand the underlying patterns of alphavirus infection, we made use of the nationwide Yellow fever testing program that has run in the country since 2003^9^. The resulting samples come from all health districts in the country and from individuals of all ages. We tested the samples of individuals who were negative for yellow fever virus infection for the presence of antibodies to three alphaviruses (ONNV, CHIKV and Mayaro virus (MAYV), which is found in South America) in a multiplex system (Magpix (MAGPIX®) Luminex platform). Further, we adapted models that explicitly consider the antibody cross reactivity between the viruses to identify a true picture of the burden from each virus^10^. Our findings allowed us to reconstruct the annual burden from each virus in the country, including where the risk is concentrated.

## Results

We tested 6324 serum samples from 186 districts for the presence of antibodies against three alphaviruses (Figure 1A-B). Of the 6324 tested samples, we had complete data from 5272 samples (age, sex, location and antibody titre data) from five years (2010, 2013, 2021, 2022 and 2023), which were used in all subsequent analyses. The mean age of people from whom the samples were collected was 18 years and 2138 (40.5%) came from females. The age and sex of the samples broadly matched the expected distribution based on Cameroon’s census data, with some oversampling of younger age groups (Figure 1C).

**Figure 1:**
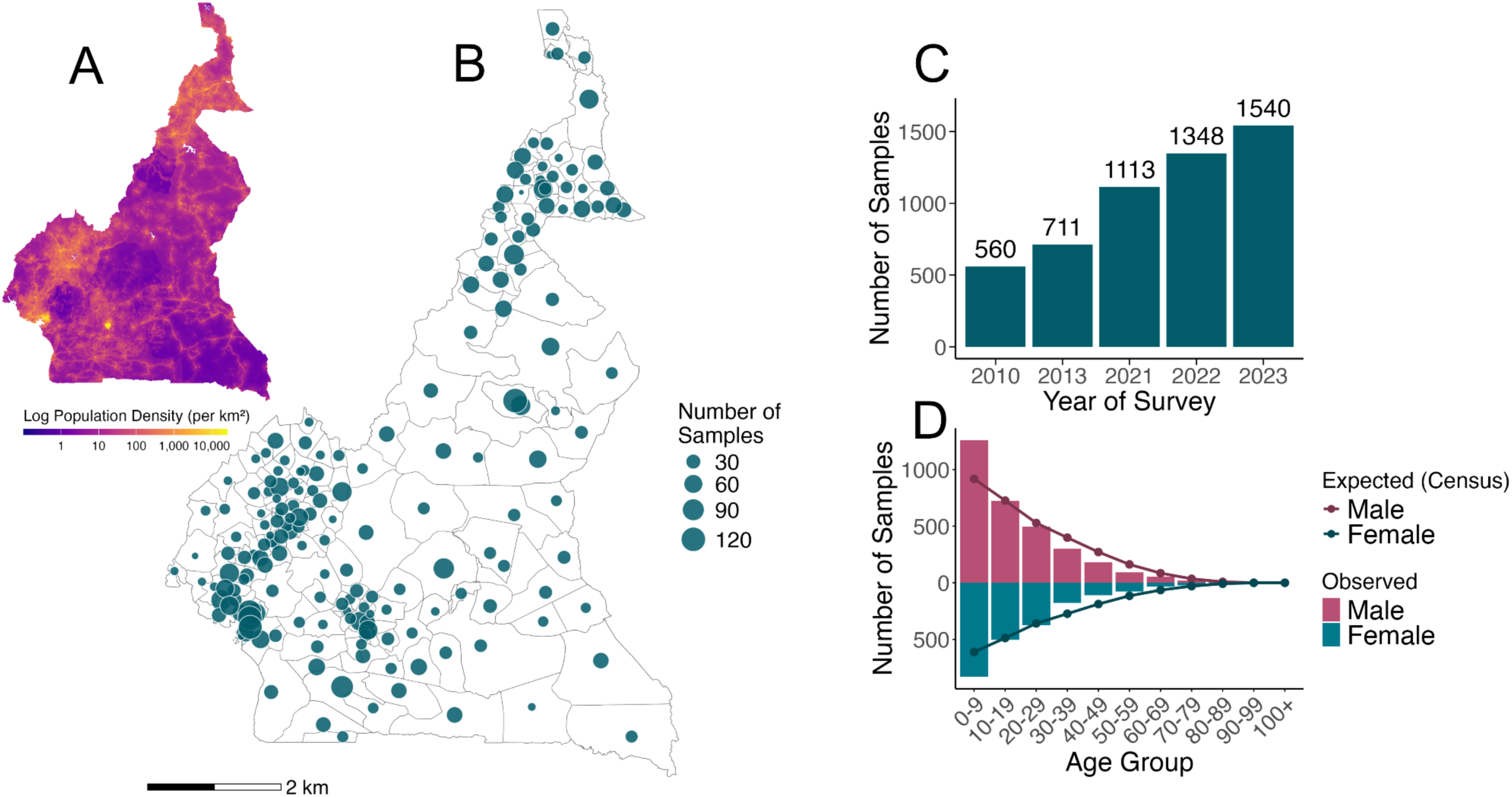
Spatial distribution, sampling coverage, and demographic characteristics of survey data from Cameroon (A) Log population density of map of Cameroon. (B) Locations of the 186 sampled districts across Cameroon, with bubble size proportional to the number of samples collected per location. (C) Total number of samples collected per survey year. (D) Age-sex distribution of survey samples compared to expected distributions derived from census data, stratified by 10-year age groups. Bars represent observed counts, lines denote census-based expected values.

To estimate the prevalence in our dataset to ONNV and CHIKV, while accounting for cross-reactivity between the viruses, we fitted a multivariate Gaussian mixture model framework. Our model was able to recover the observed distribution of titres for the three antigens (Figure 2A). We estimated that infection by ONNV resulted in a titre rise of 6.6 (95%CrI: 6.5-6.6) against the ONNV antigen, and a nearly equivalent rise of 6.5 (95%CrI: 6.3 - 6.7) against the CHIKV antigen, and the MAYV antigen (6.3, 95%CrI: 6-6.6). Infection by CHIKV resulted in a titre rise of 8.3 (95%CrI: 8.1 - 8.5) against the CHIKV antigen, a rise of 4.1 (95%CrI: 3 - 5.3) against the ONNV antigen, and a rise of 2 (95%CrI: 1.2 - 3.1) against the MAYV antigen (Figure 2B). Based on its restriction to the Western hemisphere, we assumed there was no MAYV in the population. We estimated that among the samples, there was an overall prevalence of 23.5% (95%CrI: 21.9 - 25.1) for ONNV and a prevalence of 1.7% (95%CrI: 1-2.6%) for CHIKV (Figure 2C). Had we considered the viruses independently from each other and used a two-component univariate mixture model to each response in turn, we would have estimated a prevalence of 13.4% for CHIKV, a prevalence of 23.6% for ONNV and a prevalence of 50.3% for MAYV.

**Figure 2:**
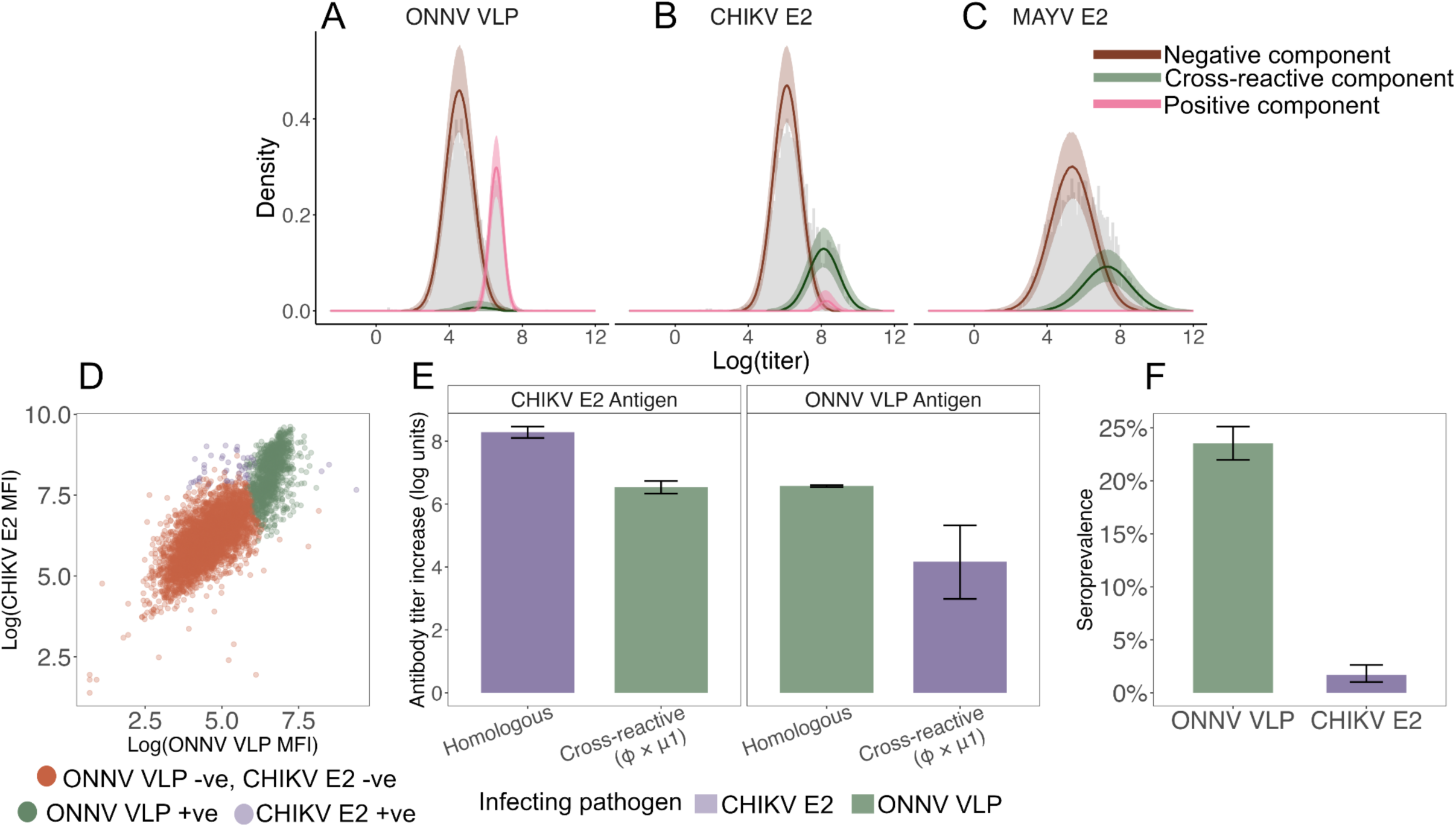
Multivariate Gaussian mixture model-based estimation of alphavirus seroprevalence and characterisation of cross-reactive antibody responses to ONNV and CHIKV. (A–C) Observed (gray bars) and reconstructed log titre distributions for (A) ONNV, (B) CHIKV, and (C) MAYV by infection status. Colored lines and shaded ribbons indicate median and 95%Crl estimates, respectively. (D) Scatter plot of log titre values for CHIKV-E2 versus ONNV-VLP antigens across individual samples, coloured by infection status. (E) Mean antibody titer increase (log units ± 95% CI) for homologous and cross-reactive responses against CHIK antigen and ONNV antigen, stratified by infecting pathogens. Cross-reactive titers shifts are shown as φ × μ1. (F) Estimated seroprevalence (%) with 95% confidence intervals for ONNV and CHIKV.

We found that the probability of a historic ONNV infection increased steadily with age across all the sampled years, consistent with long-term endemic transmission, with no evidence of differences in seroprevalence between males and females (p-value = 0.7) (Figure 3). A spatially varying catalytic model with a constant force of infection was able to recover the seropositivity by age in the data with a mean force of infection of 0.018 (95%CI: 0.015-0.022). The force of infection was spatially variable, with the highest force of infection in the east of the country (average force of infection of 0.022), and the lowest in the north-west (average force of infection of 0.011) (Table S3).

**Figure 3:**
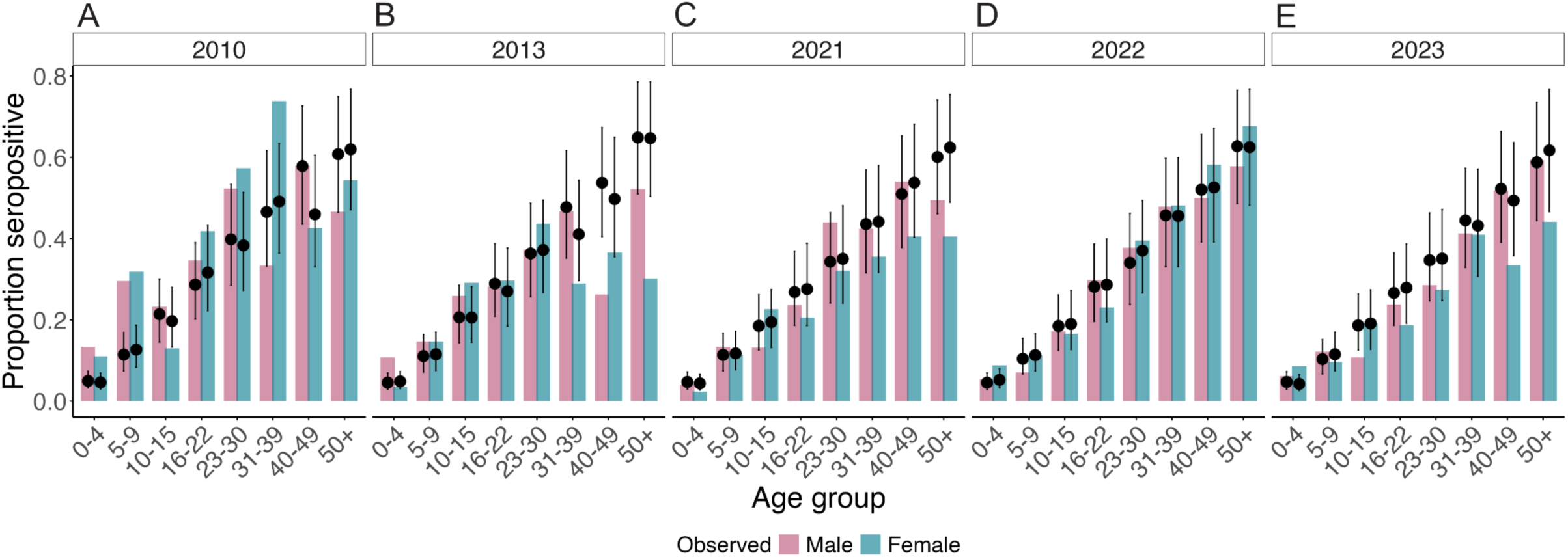
ONNV seropositivity by age and sex (A–E) Proportion seropositive by age and sex for survey years 2010, 2013, 2021, 2022, and 2023, respectively. Bars represent observed seroprevalence stratified by gender. Black dots represent median model-estimated seroprevalence and error bars (95% CI).

Using the distribution of the population and the age structure from the national census, we estimated the number of individuals that become infected each year and the total number of individuals with a history of ONNV infection. This approach accounts for differences between the location and age of the individuals in our dataset and the age and location distribution of the underlying population. We estimated that an average of 332,000 (95%CI: 289,000-382,000) individuals become infected each year and that (in 2020, the most recent year for which national gridded population data were available), 8.0 million (95%CI: 6.9-9.4m) individuals in the country had a history of ONNV infection, representing 30.4% of the population (95%CI: 26.2%-35.4%). We found that seropositivity to ONNV was strongly correlated to the presence of *An. gambaie* (p < 0.001) (Figure 4D). However, ONNV seropositivity was not correlated with the presence of *An. funestus* (p-value = 0.62), *Ae. aegypti* (p = 0.59) or *Ae. albopictus* (p = 0.25). ONNV seropositivity was negatively correlated with human population density (p-value = 0.02). The proportion of the population that had a history of CHIKV infection was not correlated with the presence of *Ae. aegypti* (p-value = 0.64), or *Ae. albopictus* (p-value = 0.7). We found that, of the 49 individuals with evidence of historic CHIKV infection from our model, 51% were located in around the two biggest cities of Douala and Yaoundé (Figure 4E).

**Figure 4:**
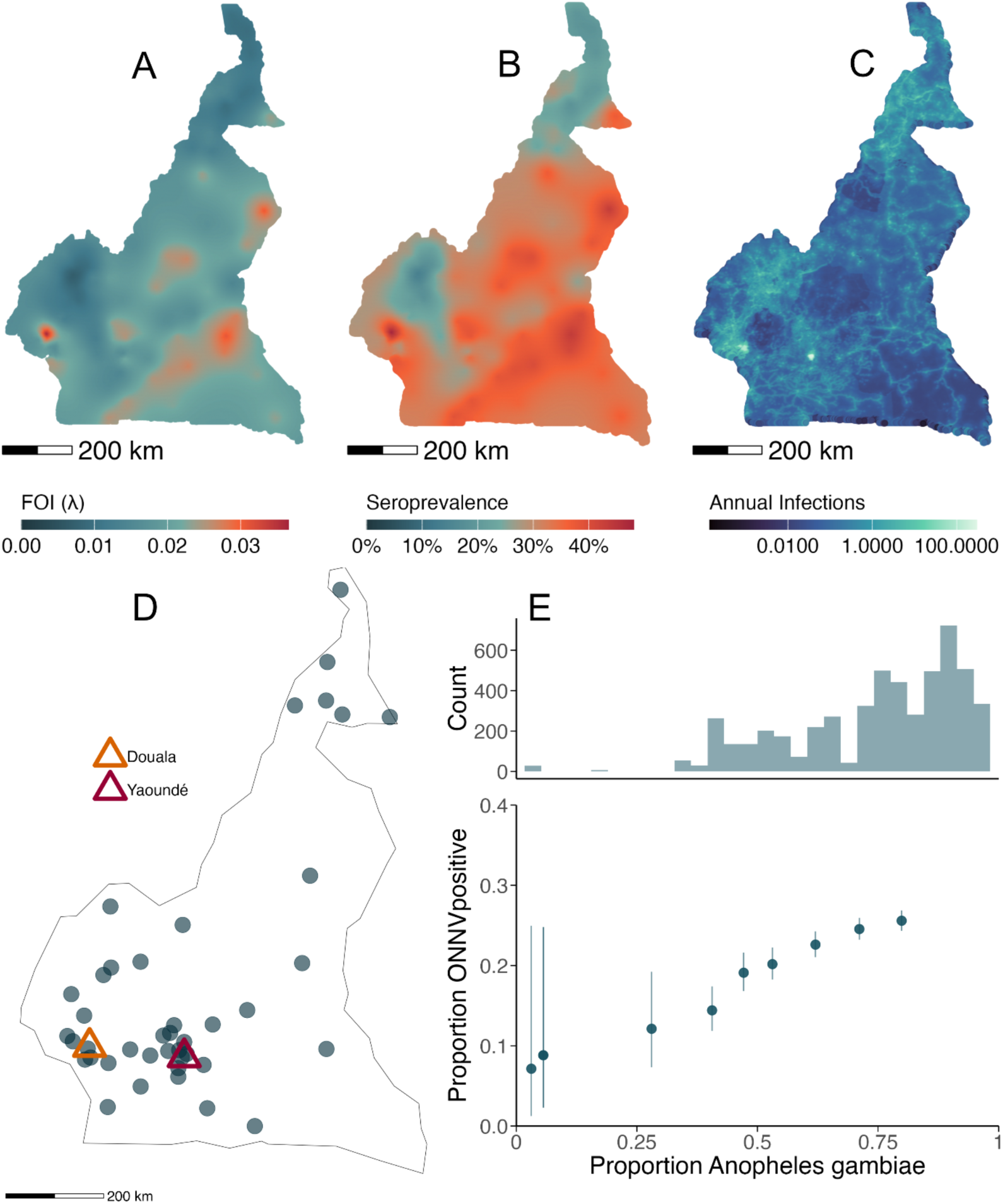
Spatial distribution of ONNV and CHIKV seropositivity in Cameroon (A) Predicted FOI of ONNV across Cameroon. (B) Predicted ONNV seroprevalence (%) across Cameroon. (C) Predicted annual ONNV infections per pixel across Cameroon (log scale). (D) Upper panel: frequency distribution of *Anopheles gambiae* suitability across sampled locations. Lower panel: Relationship between *An. gambiae* suitability and estimated ONNV seropositivity. (E) Spatial distribution of CHIKV-seropositive cases across Cameroon, with the two biggest cities (Douala and Yaoundé) highlighted.

We looked at the relationship between ONNV-positivity and malaria, using *P. falciparum* rates per person as a proxy for malaria prevalence. *P. falciparum*, the etiologic agent of malaria, is the parasite transmitted through the bite of a female *Anopheles* mosquito. We found that each one-unit increase in *P. falciparum* incidence was associated with a 17.8-fold increase in the odds of ONNV seropositivity (p<0·0001), demonstrating a positive correlation between malaria and ONNV seropositivity, likely reflecting the shared dependence of both pathogens on *Anopheles* mosquito vectors.

## Discussion

This study represents the first national estimate of the annual ONNV infection burden in any country. ONNV, historically identified in southwestern Cameroon^16^ also has evidence of circulation in neighbouring Mali and Chad^17,18^. We found that ONNV is circulating endemically across Cameroon, and has done so for many years, with an average of 332,000 infections per year. By contrast, we only found limited evidence of CHIKV circulation, consistent with short lived spatially constrained outbreaks.

The disease implications following ONNV infection are poorly quantified. There have been reports of arthralgia following ONNV infection^19,20^. In the absence of a good understanding of the relationship between ONNV infection and the development of disease, it is difficult to quantify the health burden from this endemic pathogen. Assuming that the probability of disease and death is similar to that associated with CHIKV^5,21,22^ would imply ∼166,000 acute cases, ∼ 1900 chronic arthralgia cases and ∼14 deaths per year occur in Cameroon each year from ONNV infection. There are currently no licensed vaccines for ONNV; however, there is some suggestion that live-attenuated CHIKV vaccines may provide some protection against other alphaviruses, including ONNV ^23^.

We found that, of the two candidate vector species for ONNV, *An. gambiae* was more strongly associated with ONNV infection risk. Unlike *Aedes* mosquitoes, *Anopheles* are associated with larger bodies of water, such as lakes and rivers. *Anopheles gambiae* is found throughout Africa, suggesting that ONNV may be circulating endemically across the continent. *Anopheles gambiae* is also a primary vector for malaria, and ONNV seropositivity was also strongly associated with malaria prevalence estimates, consistent with overlapping transmission ecologies (Figure S4). Other settings in Africa with a substantial malaria burden could explore the potential of a parallel ONNV burden in the same communities.

Our findings of short lived CHIKV outbreaks are consistent with prior work that has suggested that CHIKV circulates endemically in East Africa but only epidemically in the west of the continent ^5^. Chikungunya cases have been reported in Cameroon and outbreaks have occurred in Gabon, to the south, and in other surrounding countries ^2^. It is unclear why CHIKV has different ecologies in these different parts of Africa when the mosquito vectors appear widely distributed across the continent. One possibility is that enzootic transmission foci are not widespread, and therefore the risk of seeding urban epidemics varies geographically. Increased global connectivity may also increase the frequency of CHIKV introductions and subsequent outbreaks in countries such as Cameroon.

Our work highlights the need to explicitly consider serologic cross-reaction between antigenically similar viruses. Had each virus been considered independently, we would have concluded endemic circulation of CHIKV and MAYV in the population, alongside ONNV. Multiplex systems that can allow the simultaneous testing of many antigens across related pathogens are critical to explicit consideration of cross-reactivity without the need for excessive additional testing. Our work also highlights the utility of passively collected blood samples as a way of understanding the burden of infectious diseases. Nationally representative community-based serostudies have been performed, however, they are expensive and logistically complex^24^. They are also nearly always done on a one-time basis. By contrast, blood is regularly collected for many different reasons in healthcare settings.

Our study has some limitations. We did not have samples from individuals with confirmed (e.g. PCR or virus isolation) ONNV and CHIKV to validate the results of our inference. We also did not have the ability to run neutralisation testing on samples as an independent additional validation. However, the strong correlation between estimated ONNV seropositivity with the presence of ONNV vectors (and a lack of correlation with CHIKV vectors) provides strong support for our findings. A further key risk is that individuals that provide blood are not representative of the population. In this project we relied on samples collected as part of the Yellow Fever surveillance platform. The individuals who use this program do not need to pay for testing, resulting in wide availability. While individuals with suspected yellow fever tend to live in more rural environments in the absence of large urban outbreaks, individuals across the country provided our samples. We could therefore use our understanding of the demography and population distribution to nevertheless obtain national estimates of the burden. Our estimates also relied on unsupervised learning approaches.

In conclusion, despite no regular reporting of o’nyong-nyong cases in the country, we have demonstrated that ONNV is highly endemic throughout Cameroon. Now that the presence has been identified, there needs to be concerted efforts to better understand the morbidity and mortality and to identify incident cases, which will further the investment case for vaccine development against this neglected pathogen.

## Methods

### Serum samples

Stored serum samples were randomly selected from individuals who had presented to the Yellow Fever Surveillance program. The program has been running in Cameroon since 2003 and provides yellow fever testing free of charge to individuals with suspected infection ^9^. We randomly selected 6,324 samples from six years: 2010 (N=740), 2011 (N=736), 2013 (N=724), 2021 (N=1150), 2022 (N=1392), and 2023 (N=1582). All samples had been stored frozen (at -20^0^C or -80^0^C) at CPC laboratories, and represented each of the health districts in the country.

### Serological testing

Serum samples were tested using a Magpix Luminex platform at CPC laboratories with a coupled bead set provided by Institut Pasteur (Paris, France). Recombinant proteins were obtained from Native Antigen Company (Oxford, UK). The coupled bead set was performed as previously described^11^.The bead set included ONNV virus-like particles (VLP), as well as CHIKV and MAYV E2 envelope protein antigens. Samples were analysed in 96-well plates, including a blank (beads only, without serum) and a standard curve prepared from two-fold serial dilutions (1:50 to 1:102 400) of a pool of hyperimmune (HI) serum. Test samples were assayed at a final dilution of 1:400, and analysed to quantify IgG antibody levels. The 1:200 HI dilution was used to assess inter-plate variability. Plates for which the 1:200 HI control fell outside ±2 standard deviations of the expected value were excluded (13 plates; 14%). Additionally, samples with net median fluorescence intensity (MFI) values below 15 were set to NA and removed from analysis. A total of 5272 samples from 5 years were appropriate for analysis: 2010 (N=560), 2013 (N=711), 2021 (N= 1113), 2022 (N = 1384), and 2023 (N=1540).

### Estimating Seroprevalence - Multivariate Mixture Model

To determine the seroprevalence of CHIKV and ONNV in Cameroon as well as the cross reactivity between ONNV, CHIKV and MAYV viruses, we applied a semi mechanistic multivariate Gaussian mixture model. Using the model, we jointly inferred pathogen-specific prevalence estimates and between-pathogen cross-reactivity ^10^. We assumed that no MAYV was present in the population and therefore any titres to MAYV came from cross-reactivity from CHIKV or ONNV infection. The model assumed that each individual can either be infected or have never been infected with each of ONNV and CHIKV. As such, the total number of infection status combinations, c, per individual can take four possible values (0,0), (1,0), (0,1) or (1,1), where (1,0) represents having been infected by ONNV but not by CHIKV. Then, the proportion of the study population that falls into each combination, denoted as **θ**_c_, is the conditional probability of having an infection combination, given the pathogen-specific population prevalence, π_P_. For example, for infection status combination where individuals only have been infected with ONNV (c_1_ = (1, 0)), the proportion of the population with infection status c_1_ is: **θ**_c_ = (π_ONNV_) (1- π_CHIK_).

For each infection status combination, a *P*-dimensional Gaussian component was defined by a mean vector **μ*_c_*** and covariance matrix **σ_c_**, which captures the expected antibody titre shifts due to homologous and cross-reactive infection. For each circulating virus, the mean baseline (uninfected) titre value was defined as μ_●-0_ with standard deviation σ_●-0_. Upon infection with only one pathogen, for example ONNV, (c_1_ = (1, 0)), individuals experience a rise from their baseline antibody titre against ONNV, defined as μ_ONNV-0_ + μ_ONNV-1_ with standard deviation σ_ONNV-1_ where μ_1_ is the increase in antibody titre due to homologous infection. Individuals also experience a relative increase in antibody titre against CHIKV after infection with ONNV defined as μ_CHIKV-0_ + (ϕ_ONNV → CHIKV_) μ_ONNV-1_ where ϕ is the relative cross-reactive increase in antibody titres and ϕ_ONNV → CHIKV_ is the increase in antibodies raised against CHIKV after infection with ONNV.

For each individual *i*, the likelihood contribution was computed by summing over all possible infection status combinations, where each combination was weighted by its prevalence, θ_c_, in the population. The probability of observing individual *i’s* antibody titre profile, *x_i_* given infection status combination c was modelled as a multivariate normal distribution with mean vector **μ**_c_ and covariance matrix **Σ**_c_, denoted as N(**x**_i_ | **μ**_c_, **Σ**_c_). The full model likelihood was then calculated as the sum of the log probabilities across individuals, *i*, and across all possible infection status combinations, *c*, shown in Eq. 1.

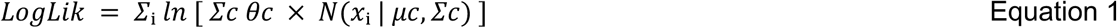

### Parameter Estimation

We fitted the model in a Bayesian framework with Hamiltonian Monte Carlo No-U-Turn sampling using cmdStanR^12^. Three chains for 3000 iterations in addition to 1000 warm-up samples were run. Parameter priors, which were assumed to be constant across pathogens, are shown in Table S1. Median and 95%CrI estimates were calculated for each parameter.

### Estimating Seroprevalence - 1D Mixture Model

To illustrate the potential for misclassification when cross-reactivity between pathogens is not accounted for, we also fitted univariate mixture models, where each virus is considered separately. For each pathogen we fitted a two-component finite mixture model, assuming normally distributed log transformed antibody titres, using the *mixfit* package in R to classify individuals as seronegative and seropositive. Prevalence estimates from these univariate models were then compared to those from the multivariate mixture model described above.

### Spatial Prediction across Cameroon

To explore the variability of ONNV prevalence across Cameroon, we applied a spatially explicitly catalytic model with a Bayesian framework with a Matern spatial correlation structure, implemented using INLA^13^. We used the geocoded survey data, aggregated at the district level. District names in the survey data were matched using administrative shapefiles. Districts absent from shapefiles were resolved by geocoding their administrative coordinates, and spatially assigning each to its nearest shapefile polygon, reducing the total districts from 198 to 186. Population-weighted centroids, derived using population data from WorldPop^14^, were then calculated for each district.

We first assigned individuals a binary serostatus to ONNV by selecting the infection state with the highest posterior probability under the multivariate Gaussian mixture model. Subsequently, we fit the serostatus of each individual to a binomial model with a *cloglog* link function. We include log(age) as an offset, and place a prediction grid over the country, derived from WorldPop population data, allowing us to directly estimate spatially varying force of infection (FOI) (or “hazard”) for each 100m x 100m prediction location, *x* ^14^. Using the location specific FOI, we estimate the location specific proportion seropositive at age *a*, given by the following expression:

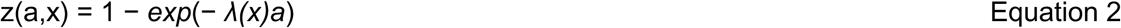

where λ(x) is the location-specific estimated force of infection and *a* is age.

We used census data to obtain the age-stratified population structure of Cameroon ^15^. The overall seroprevalence at each location was then calculated as the average seroprevalence within each age interval, weighted by the proportion of the total population within each age group.

To estimate the annual number of new infections, we combined the location-specific FOI estimates with population data, obtained from WorldPop ^14^. For each prediction location and age group, we calculated the number of susceptible individuals as the product of the total population in that age group {N(a,x)} and the average proportion susceptible within the age interval. For each location *x*, the proportion susceptible at age *a* is given by S(a,x) = exp(−λ(x)a), which we integrated over each age interval (0-4, 5-9, …, 95-99, 100+ years) to obtain the average susceptibility for that age group. Annual infections in each age group and location were then estimated as:

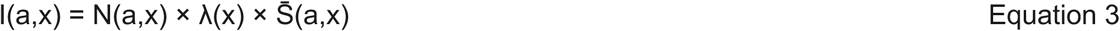

where I(a,x) is the number of annual infections, N(a,x) is the population in age group *a* at location x, λ(x) is the FOI at location x, and Ŝ(a,x) is the average proportion susceptible in age group a at location x. Total annual infections were estimated by summing across all age groups and grid cells.

### Seropositivity correlation with mosquito vectors

We explored the association between individual estimated seropositivity and a set of ecological covariates. We obtained data on the predicted habitat suitability for the primary mosquito vectors of ONNV and CHIKV; *Anopheles funestus* and *Anopheles. gambiae* for ONNV^8^, and *Aedes aegypti* and *Aedes. albopictus* for CHIKV^7^. We additionally examined associations with log-transformed population density^14^ and *Plasmodium falciparum* parasite rate^8^, the latter included as a proxy for malaria prevalence given the shared dependence of ONNV and malaria on Anopheles vectors. For each covariate, we assigned each individual in our dataset into a bin based on the prevalence of the covariate in the home location of that individual. For each bin, we then calculated the mean proportion seropositive. To formally test each association, we also fitted a binomial logistic regression model with binary serostatus (based on a cut-off of 50% probability of positivity) as the outcome and the covariate of interest as the predictor and estimated the direction and strength of each relationship.

## Data Availability

All data used in this study is available online at https://github.com/anchitapuri/cameroon_chik_onnv

## Acknowledgements

This study received funding from the coordinating center of the NIH/NIAID/Centers for Research in Emerging Infectious Diseases (CREID), including pilot award number 1U01AI151378), the West African Center for Emerging Infectious Diseases (WAC-EID) grant Number 1U01AI151801, the Cambridge-Africa ALBORADA Research fund and the Wellcome Trust (number 228185/Z/23/Z). NNA is supported by the European Union (grant no. DCI-PANAF/2020/420-028) through the African Research Initiative for Scientific Excellence (ARISE program). MW was supported by the French government’s “Integrative Biology of Emerging Infectious Diseases” (Investissement d’Avenir grant ANR-10-LABX-62-IBEID) and INCEPTION (Investissement d’Avenir grant ANR-16-CONV-0005) programs.

## Data and code availability statement

*All the data and code used for the analysis presented here is publicly available in the following GitHub repository: https://github.com/anchitapuri/cameroon_chik_onnv*

## Role of the funding source

*The funder of the study had no role in study design, data collection, data analysis, data interpretation, or writing of the report*.

## Supplementary materials

**Table 1:** Multivariate mixture model parameter priors. Parameter priors and limits used in model fitting.

| Parameter | Parameter description | Prior |
| --- | --- | --- |
| $\pi$ | Prevalence | beta (1,5) |
| $\mu_0$ | Mean of negative titres | normal (5,0.5) |
| $\mu_1$ | Mean increase of positive titres | normal (2.5,0.1) |
| $\sigma_0$ | SD of negative titres | normal (1,0.1) |
| $\sigma_1$ | SD of positive titres | normal (0.3, 0.05) |
| $\phi$ | Cross reactivity | lognormal (log (0.3),0.3) |
| $\rho$ | Correlation of negative titres | beta (3,7) |

**Table 2:** Comparison of the Full model (ONNV and CHIKV samples) with the ONNV only model (including the 920 samples missing CHIKV measurements). Median parameter estimates with 95% CI and FOI estimates for both models. For Model 1, which assumes the presence of ONNV and CHIK (and absence of MAYV), 920 samples with missing CHIK data were excluded. To assess the impact of this exclusion, we fitted two versions of Model 2: one using only ONNV titre data, and one incorporating the full dataset, including the 920 samples missing CHIK measurements but complete for ONNV and MAYV. Median parameter estimates were broadly comparable across both versions, and spatially varying FOI estimates were derived for each.

| **Multivariate Mixture Model Parameter estimates** |  |  |
| --- | --- | --- |
| Parameter | **Model 1 (Full Model:**<br>**ONNV +ve, CHIKV +ve)** | **Model 2 (ONNV Only**<br>**Model)** |
| $\pi_{\text{ONNV}}$ | 0.23 (95%CI:0.22 - 0.25) | 0.23 (95%CI:0.21 - 0.24) |
| $\mu_{0\text{ONNV}}$ | 4.54 (95%CI: 4.51 - 4.57) | 4.64 (95%CI: 4.61 - 4.67) |
| $\mu_{1\text{ONNV}}$ | 6.57 (95%CI: 6.54 - 6.60) | 6.65 (95%CI: 6.62 - 6.68) |
| $\phi_{\text{ONNV} \rightarrow \text{MAYV}}$ | 0.97(95%CI: 0.92 - 1.0) | 1.0 (95%CI: 0.97 -1.05) |
| **Spatially varying FOI** |  |  |
|  | **Model 1 (Full Model:**<br>**ONNV +ve, CHIKV +ve)** | **Model 2 (ONNV Only**<br>**Model)** |
| FOI | 0.018 (95% CI: 0.015 - 0.02) | 0.017 (95% CI: 0.014 - 0.02) |

**Table 3:** FOI, Seropositivity and Annual Infections estimates by region.

| **Region** | **Region Population** | **Population Weighted FOI estimate** | **Population Weighted seropositivity estimate** | **Annual Infections** |
| --- | --- | --- | --- | --- |
| Adamawa | 1434657 | 0.0197 | 0.32 | 17233 |
| Centre | 5450965 | 0.0194 | 0.32 | 71826 |
| East | 1207469 | 0.022 | 0.35 | 14327 |
| Far North | 4608223 | 0.013 | 0.24 | 43315 |
| Littoral | 4073615 | 0.018 | 0.30 | 49880 |
| North | 3082097 | 0.016 | 0.28 | 32889 |
| North-West | 2570428 | 0.011 | 0.199 | 18295 |
| West | 2046142 | 0.012 | 0.22 | 18475 |
| South | 1154882 | 0.021 | 0.34 | 12716 |
| South-West | 2087437 | 0.019 | 0.31 | 23092 |

**Supplementary Figure 1.**
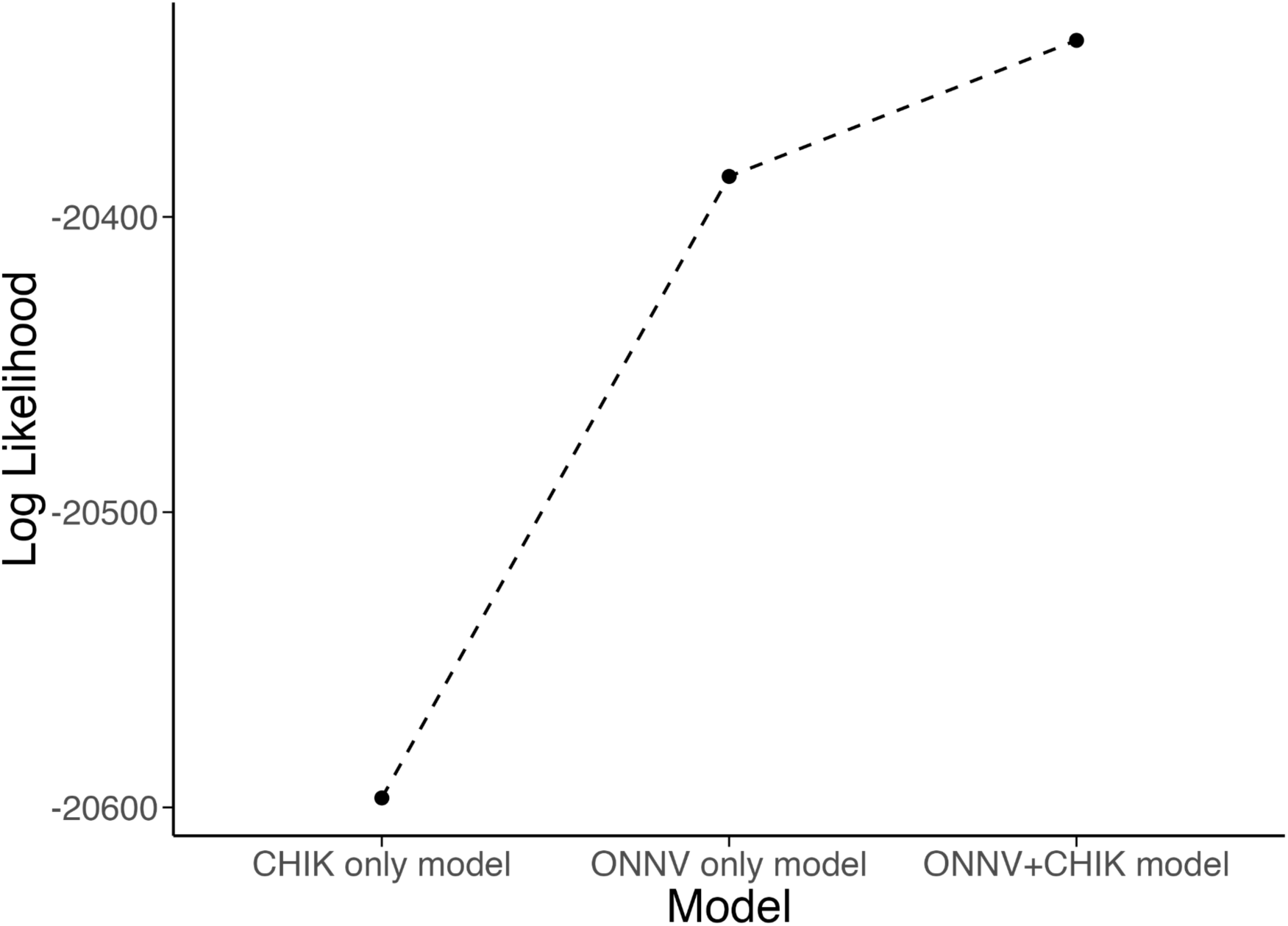
Model fit comparison by log-likelihood across three models (CHIKV-only, ONNV-only, and ONNV+CHIKV). The combined ONNV+CHIKV model achieved the highest log-likelihood, supporting its selection as the final model.

**Supplementary Figure 2:**
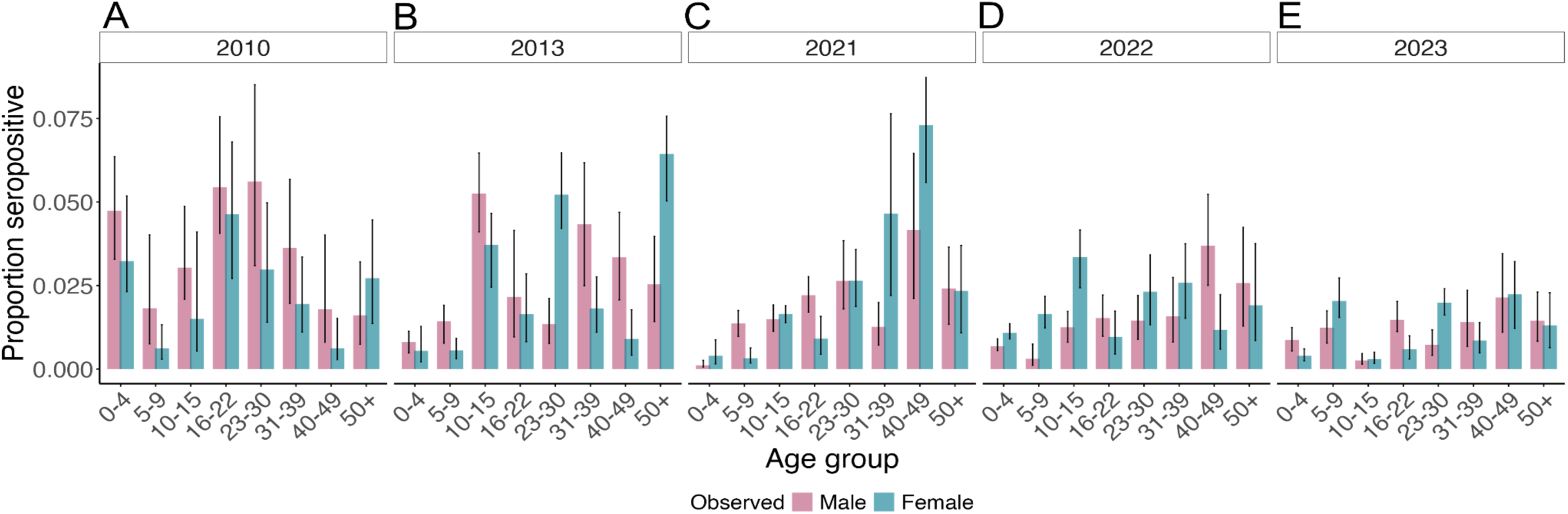
CHIKV seropositivity by age and sex (A–E) Proportion seropositive by age and sex for survey years 2010, 2013, 2021, 2022, and 2023, respectively. Bars represent median seroprevalence estimated by multivariate mixture model, with 95%CrI, stratified by gender.

**Supplementary Figure 3:**
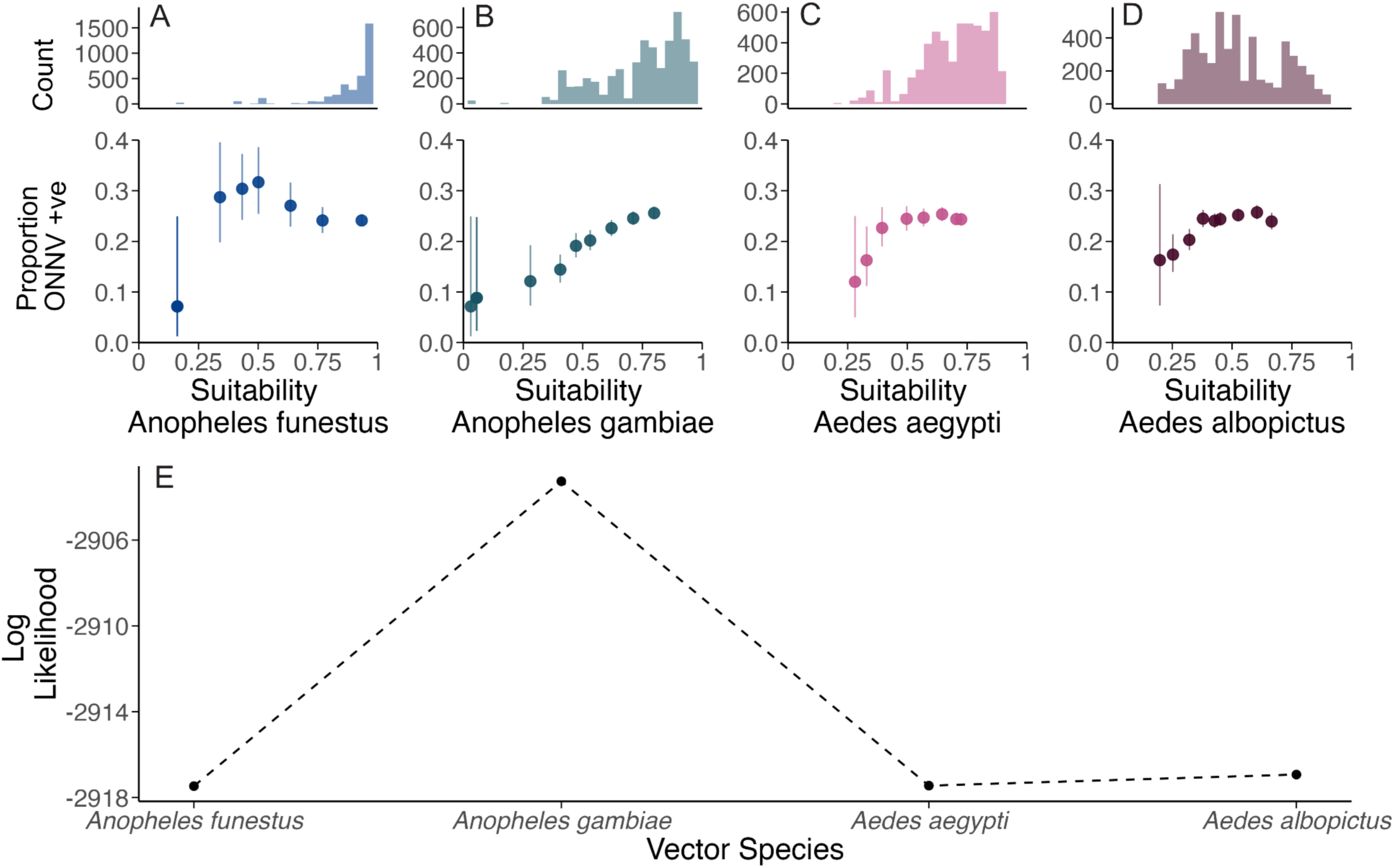
(A–D) For each of four mosquito vectors: Anopheles funestus (blue), Anopheles gambiae (teal), Aedes aegypti (pink), and Aedes albopictus (mauve), upper panels show the frequency distribution of suitability across sampled locations, and lower panels show the relationship between ONNV seropositivity and vector suitability. **(E)** Log-likelihood comparison across four single-vector models, each incorporating the suitability of one candidate vector species as a covariate. Anopheles gambiae suitability yielded the highest log-likelihood, indicating the best model fit and supporting that Anopheles gambiae is associated with ONNV risk.

**Supplementary Figure 4:**
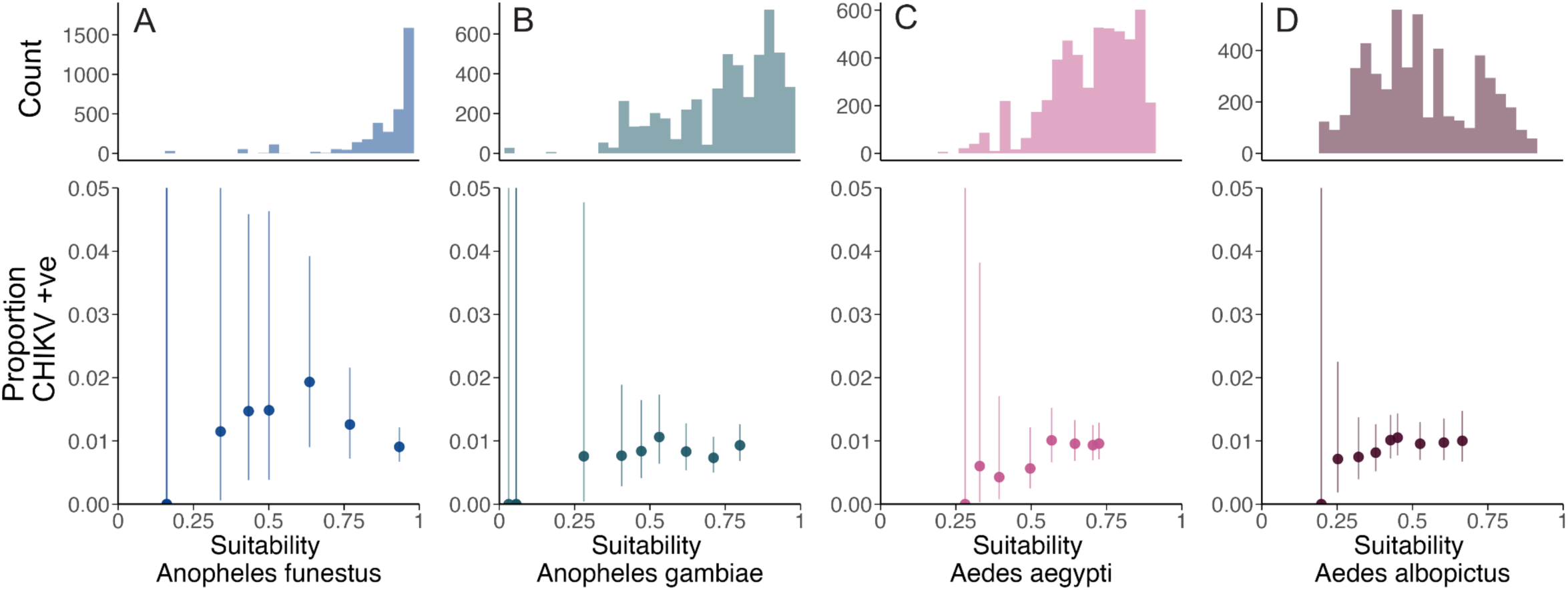
(A–D) For each of four mosquito vectors: Anopheles funestus (blue), Anopheles gambiae (teal), Aedes aegypti (pink), and Aedes albopictus (mauve), upper panels show the frequency distribution of suitability across sampled locations, and lower panels show the relationship between CHIK seropositivity and vector suitability.

**Supplementary Figure 5:**
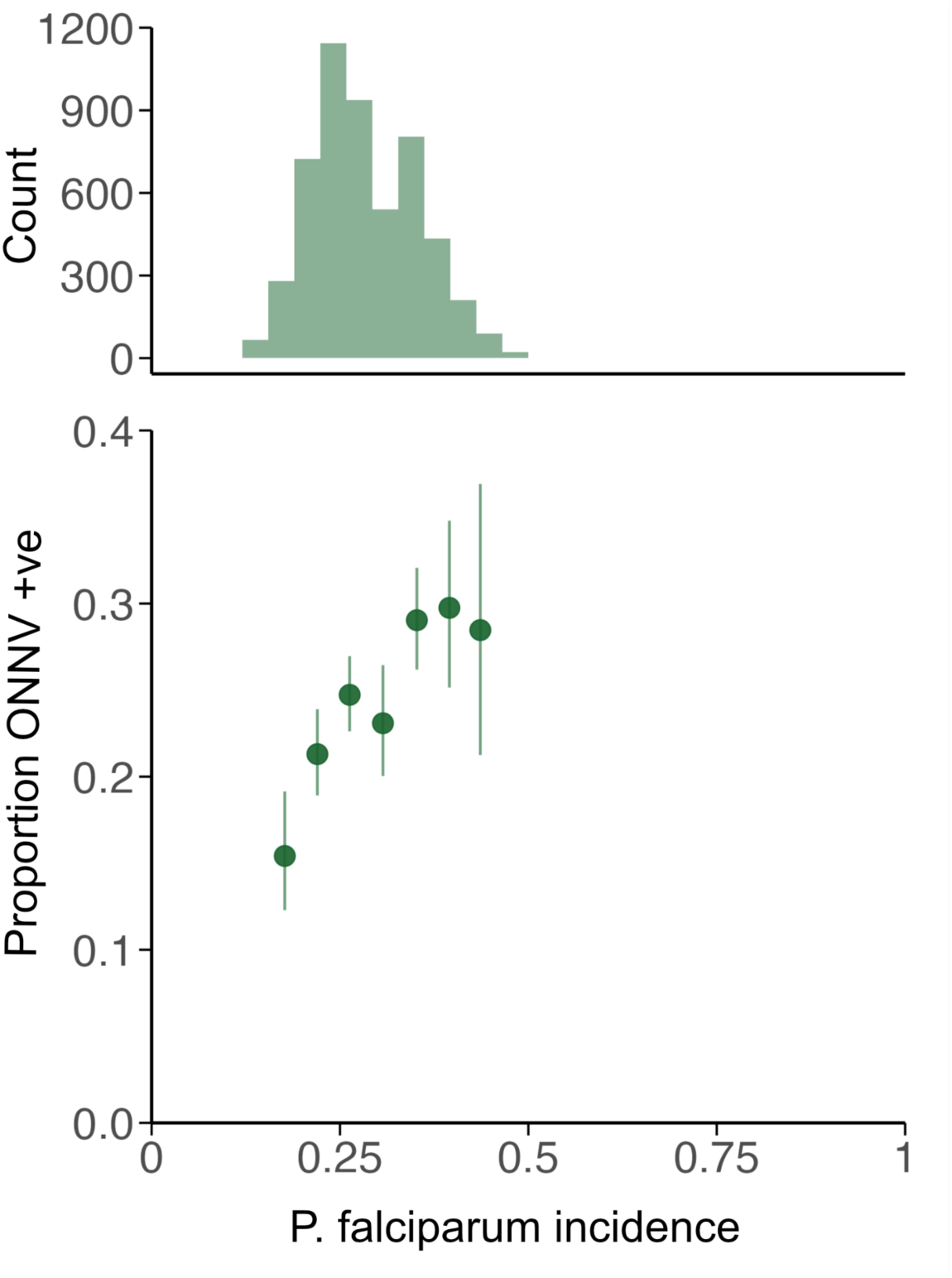
Upper panel: frequency distribution of *P. falciparum* incidence across sampled locations. Lower panel: Relationship between ONNV seropositivity and *P. falciparum* incidence.

## Notes

### Competing Interest Statement

The authors have declared no competing interest.

### Author Declarations

Centre Pasteur du Cameroon (CPC) Ethics Review Board of Centre Pasteur du Cameroon (CPC) gave ethical approval for this work.

